# FACTORS CONTRIBUTING TO TUBERCULOSIS TREATMENT DEFAULT IN KALINGALINGA COMPOUND IN LUSAKA ZAMBIA

**DOI:** 10.1101/2025.08.20.25334134

**Authors:** Luyando Mainza, Chipampe Lombe

## Abstract

Treatment default in tuberculosis care undermines disease control efforts, contributing to ongoing transmission, drug resistance, and adverse health outcomes. Zambia, like many high-burden countries, continues to face challenges with patient adherence to tuberculosis (TB) treatment, especially in urban informal settlements. Despite the national TB program’s efforts, limited research has specifically explored the causes of treatment default in these settings. This study was conducted in Kalingalinga Compound, Lusaka, to identify and understand the factors contributing to default from TB treatment, and to inform the development of more effective, context-specific interventions.

**Methods:** A cross-sectional study was conducted among 180 tuberculosis patients who had defaulted on treatment in Kalingalinga Compound, Lusaka. Participants were selected using stratified random sampling based on age, gender, and socio-economic status to ensure adequate representation. Data were collected through structured questionnaires covering socio-demographic characteristics, patient-related factors, and health system influences. The data were analysed using Statistical Package for the Social Sciences (SPSS) Version 26. Descriptive statistics were used to summarise the data, while inferential statistics including chi-square tests and logistic regression were applied to determine associations and predictors of treatment default, with results reported at a 95% confidence interval.

**Results:** The study found that male gender (AOR = 1.86, p = 0.041), age group 30–49 years (AOR = 2.74, p = 0.006), and low education levels (AOR = 3.12, p = 0.004) were significant demographic predictors of TB treatment default. Patient-related factors such as fear of stigma (AOR = 4.21, p < 0.001), substance abuse (AOR = 2.85, p = 0.004), and inadequate knowledge of TB (AOR = 3.17, p = 0.004) were strongly associated with default. Health system deficiencies, including inadequate counselling services (AOR = 5.12, p < 0.001), inconsistent medication availability (AOR = 3.89, p < 0.001), and long distances to health facilities (AOR = 3.44, p < 0.001), significantly contributed to non-adherence.

**Conclusion:** The findings highlight the multifaceted nature of TB treatment default in Kalingalinga Compound. Interventions should address gender and age-specific barriers, enhance patient knowledge, reduce stigma, and improve healthcare system delivery. Holistic and locally tailored strategies are necessary to improve adherence and reduce the TB burden in Zambia.

## Introduction

Tuberculosis (TB) remains a major public health challenge in Zambia, which is classified among the 30 high-burden TB countries globally. In 2021, the country reported over 500,000 TB cases, with a significant proportion arising from urban informal settlements such as Kalingalinga Compound in Lusaka [1]. Despite the implementation of the Directly Observed Treatment, Short-course (DOTS) strategy and the expansion of TB services in public health facilities, treatment default continues to hinder TB control efforts. This results in prolonged infectiousness, increased mortality, and the emergence of drug-resistant TB strains [2].

Zambia also faces a dual epidemic of TB and HIV, with co-infection increasing the risk of developing active TB and complicating both diagnosis and management [3]. In high-density, low-income areas like Kalingalinga, the burden of TB is worsened by factors such as poverty, overcrowding, poor housing, malnutrition, alcohol and substance use, and limited access to healthcare services [4]. Although TB treatment is provided free of charge, treatment adherence remains suboptimal. While national TB program reports indicate relatively low default rates, evidence from urban informal settlements suggests higher rates of treatment default, often driven by socio-economic constraints and systemic health barriers [5]. Contributing factors include lack of transportation, poor patient follow-up, limited health literacy, stigma, and inconsistent drug supply [6].

Although prior research has explored general factors influencing TB non-adherence in Zambia, few studies have focused specifically on treatment default in high-risk communities like Kalingalinga. Additionally, relying solely on quantitative data may overlook critical patient-level and contextual nuances that influence default behaviour. A better understanding of the interaction between individual, socio-economic, and health system factors is essential to inform targeted interventions. This study aimed to identify and analyse the key factors associated with TB treatment default among patients in Kalingalinga Compound. The goal is to generate locally relevant evidence to inform more effective strategies for improving adherence and advancing TB control efforts in Zambia.

## Materials and Methods

### Ethical Considerations

This study was conducted in full compliance with ethical research standards. Ethical approval was granted by the University of Lusaka under reference number FWA00033228-032(08)/(08)/(2024). Additionally, authorization to conduct the research was obtained from the National Health Research Authority (NHRA) of Zambia, as confirmed in the approval letter dated 2nd September 2024.Participants were thoroughly informed about the study’s purpose, procedures, potential benefits, and risks. Written informed consent was obtained from each participant prior to their involvement. Measures to ensure confidentiality included anonymizing personal identifiers and securely storing data, with access limited to the research team. The study adhered to core ethical principles of autonomy, beneficence, and justice, safeguarding the rights and welfare of all participants throughout the research process.

### Study setting and population

The study was conducted in Kalingalinga Compound, a densely populated urban settlement in Lusaka, Zambia. The area is characterised by high tuberculosis (TB) prevalence, socio-economic hardships, and limited access to healthcare services, making it a recognised TB hotspot. Local healthcare facilities in Kalingalinga provide TB diagnosis and treatment services to the community, which comprises a mix of low-income households and informal sector workers. This study focused on patients diagnosed with drug-susceptible TB who subsequently defaulted on treatment. Patients are diagnosed and initiated on treatment following Zambia’s National TB Control Program guidelines, with first-line regimens administered through community-based clinics. Defaulters were identified from clinic records of registered TB patients who had interrupted their treatment after initiation.

The inclusion criteria required participants to be residents of Kalingalinga, diagnosed with TB, started on treatment, and recorded as having defaulted. Patients who were still on treatment or had completed treatment were excluded. Treatment default was defined as missing scheduled doses for a sufficient period to be classified by healthcare providers as non-adherent to therapy. A total of 207 patients who had interrupted TB treatment were identified in clinic records. Using Yamane’s formula, with a 95% confidence interval and 5% margin of error, a final sample size of 180 participants was calculated. Patients were selected using stratified random sampling based on age, gender, and socio-economic status to ensure diversity and representativeness across the population. Participants were contacted through clinic databases and included only if they had previously consented to be approached for research purposes.

### Data Collection

Data were collected using structured questionnaires administered by trained data collectors. The questionnaires were designed to capture comprehensive information relevant to the study objectives, with sections covering socio-demographic characteristics, patient-related factors, and health system issues. Data collectors received thorough training to ensure consistent administration of the questionnaires and to minimise interviewer bias. Participants were given clear explanations of each question to promote accurate and complete responses. The structured format of the questionnaire facilitated standardisation across respondents, thereby supporting efficient data entry and analysis.

The primary data collection instrument was a structured questionnaire that had been pre-tested to ensure reliability and validity. The pre-testing process involved administering the questionnaire to a small group of TB patients outside the study area, enabling the identification and correction of any ambiguities or inconsistencies. The final version of the questionnaire consisted of three main sections. The first section captured socio-demographic information such as age, gender, education level, and income. The second section focused on health system factors, including access to healthcare facilities, quality of care, and availability of support services. The third section explored patient-related factors, including knowledge about TB, attitudes towards treatment, and socio-economic conditions. This comprehensive structure ensured that all relevant aspects influencing TB treatment default were adequately addressed

### Data Analysis

Data were analysed using the Statistical Package for the Social Sciences (SPSS), version 26. Descriptive statistics, including frequencies, percentages, means, and standard deviations, were used to summarise socio-demographic characteristics and other key variables relevant to the study. Chi-square tests were conducted to explore associations between categorical variables and TB treatment default. Binary logistic regression was then performed to assess the strength and direction of these associations. The regression model included variables across three domains: socio-demographic factors (such as age, gender, and education), patient-related factors (such as fear of stigma, substance use, and TB knowledge), and health system factors (such as distance to healthcare facilities, availability of medication, and quality of counselling services).

Variables that showed significant associations in bivariate analyses were entered into a multivariate logistic regression model. Adjusted odds ratios (AORs) with 95% confidence intervals (CIs) were calculated to identify independent predictors of TB treatment default. A p-value of less than 0.05 was considered statistically significant. This analytical approach enabled identification of key factors influencing TB treatment default, providing a robust evidence base for subsequent discussion and policy recommendations.

## RESULTS

### Demographic Characteristics

Out of 180 respondents, 112 (62.2%) were male and 68 (37.8%) were female. Logistic regression analysis showed that males were significantly more likely to default on TB treatment than females, with an Adjusted Odds Ratio (AOR) of 1.86, 95% Confidence Interval (CI): 1.02–3.38, and a p-value of 0.041. In terms of age, 50 respondents (27.8%) were aged 18–29, 100 (55.6%) were aged 30–49, and 30 (16.7%) were aged 50 and above. The age group 30–49 had significantly higher odds of defaulting compared to the ≥50 age group, with an AOR of 2.74, 95% CI: 1.34–5.61, and a p-value of 0.006. No significant association was observed for the 18–29 age group. Regarding education, 20 respondents (11.1%) had no formal education, 80 (44.4%) had primary education, 60 (33.3%) had secondary education, and 20 (11.1%) had tertiary education. Respondents with no education had over three times the odds of defaulting (AOR = 3.12, CI: 1.45–6.71, p = 0.004), and those with primary education were also significantly more likely to default (AOR = 2.65, CI: 1.33–5.28, p = 0.006) compared to those with tertiary education. Secondary education was not significantly associated with default (p = 0.298). These results indicate that being male, aged 30–49, and having a lower level of education are significant predictors of TB treatment default.

**Table 1:** summarizes the demographic characteristics of the respondents and their association with TB treatment default (n=180)

| Variables | Frequency (n) | Percentage (%) | AOR | CI | p-value |
| --- | --- | --- | --- | --- | --- |
| **Gender** |  |  |  |  |  |
| Male | 112 | 62.2 | 1.86 | 1.02–3.38 | 0.041* |
| Female | 68 | 37.8 | Reference |  |  |
| **Age Group (Years)** |  |  |  |  |  |
| 18–29 | 50 | 27.8 | 1.21 | 0.56–2.60 | 0.621 |
| 30–49 | 100 | 55.6 | 2.74 | 1.34–5.61 | 0.006* |
| ≥50 | 30 | 16.7 | Reference |  |  |
| **Education Level** |  |  |  |  |  |
| None | 20 | 11.1 | 3.12 | 1.45–6.71 | 0.004* |
| Primary | 80 | 44.4 | 2.65 | 1.33–5.28 | 0.006* |
| Secondary | 60 | 33.3 | 1.48 | 0.71–3.10 | 0.298 |
| Tertiary | 20 | 11.1 | Reference |  |  |

\*= Significant finding, AOR = Adjusted Odds Ratio, CI = 95% Confidence Interval

### Patient-Related Factors Influencing TB Treatment Default

Among the 180 respondents analysed, key patient-related factors were found to significantly contribute to TB treatment default. Fear of stigma emerged as the most prominent predictor, with 140 individuals (93.3%) who reported experiencing stigma defaulting on treatment. These patients had over four times higher odds of defaulting compared to those who did not report stigma (AOR = 4.21, 95% CI: 2.01–8.81, p < 0.001). Substance abuse was another significant factor, reported by 100 respondents (83.3%), and was associated with nearly three times higher odds of defaulting (AOR = 2.85, 95% CI: 1.39–5.86, p = 0.004). Additionally, inadequate knowledge of TB was reported by 70 respondents (87.5%), and was significantly linked to treatment default (AOR = 3.17, 95% CI: 1.44–6.98, p = 0.004). These findings highlight the critical influence of psychosocial and informational barriers particularly stigma, substance abuse, and poor knowledge on TB treatment adherence.

**Table 2:** Patient-Related Factors and TB Treatment Default (n=180)

| Variables | Defaulted<br>(n, %) | AOR | 95% CI | p-value |
| --- | --- | --- | --- | --- |
| Fear of Stigma (Yes vs. No) | 140 (93.3%) | 4.21 | 2.01–8.81 | <0.001* |
| Fear of Stigma (Yes vs. No) | 100 (83.3%) | 2.85 | 1.39–5.86 | 0.004* |
| Knowledge of TB (Inadequate vs. Adequate) | 70 (87.5%) | 3.17 | 1.44–6.98 | 0.004* |

\*= Significant finding, AOR = Adjusted Odds Ratio, CI = 95% Confidence Interval

### Health System Factors Influencing TB Treatment Default

Health system-related barriers were found to significantly influence TB treatment default among respondents. A total of 120 individuals (85.7%) who reported living far from a health facility defaulted on treatment, with these respondents being over three times more likely to default compared to those living nearby (AOR = 3.44, 95% CI: 1.67–7.07, p < 0.001). Inconsistent availability of medication was also a major factor, with 100 respondents (90.9%) defaulting, and nearly four times higher odds of defaulting observed among this group (AOR = 3.89, 95% CI: 1.92–7.87, p < 0.001). Additionally, inadequate counselling services were associated with the highest likelihood of default, as 130 respondents (92.9%) affected by this issue were over five times more likely to default on treatment compared to those who received adequate counselling (AOR = 5.12, 95% CI: 2.45–10.67, p < 0.001).

**Table 3:** Health system-related barriers are summarized (n=180)

| Variables | Defaulted<br>(n, %) | AOR | 95% CI | p-value |
| --- | --- | --- | --- | --- |
| Distance to Health Facility (Far vs. Near) | 120<br>(85.7%) | 3.44 | 1.67–7.07 | <0.001* |
| Availability of Medication (Inconsistent vs. Consistent) | 100<br>(90.9%) | 3.89 | 1.92–7.87 | <0.001* |
| Counselling Services (Inadequate vs. Adequate) | 130<br>(92.9%) | 5.12 | 2.45–10.67 | <0.001* |

\*= Significant finding, AOR = Adjusted Odds Ratio, CI = 95% Confidence Interval

### Discussion

The findings of this study demonstrate that tuberculosis (TB) treatment default in Kalingalinga Compound is driven by an interrelated set of demographics, individual, and health system factors. Males were significantly more likely to default compared to females. This may be attributed to gendered socioeconomic roles in Zambia where men, as primary breadwinners, may prioritize work and income over consistent healthcare attendance. Occupational demands, loss of income during clinic visits, and limited flexibility in health service hours likely contribute to male default, reinforcing findings from similar studies in Tanzania and Uganda [7,8]. Conversely, women may be more consistent in seeking care due to maternal and familial responsibilities and possibly greater exposure to community health programs targeting maternal and child health.

Default was most prevalent among individuals aged 30–49 years, a demographic considered economically productive and socially mobile. The competing demands of employment, family care, and societal expectations in this age bracket may limit the perceived priority of TB treatment, especially in cases where symptom relief occurs early in the course of treatment. The social stigma attached to TB can also be particularly distressing for individuals in this age group, where productivity and social perception are critically valued. This aligns with studies in urban Ethiopia and South Africa that link TB stigma with reduced adherence, particularly among working-age adults [9,10].

Educational attainment showed a significant association with treatment adherence. Participants with no formal education or only primary schooling had a higher likelihood of defaulting. Low education levels correlate with poor understanding of disease transmission, treatment duration, and consequences of default. These patients are also more susceptible to misinformation and traditional health beliefs that may discourage the use of biomedicine or suggest stopping treatment after symptomatic improvement. This is consistent with findings in multiple African settings where health literacy plays a critical role in TB outcomes [11,9]. It suggests a need for culturally appropriate, simplified, and repetitive educational messaging delivered through trusted community structures such as churches, traditional leaders, and local health volunteers.

Patient-related factors were found to be some of the strongest predictors of treatment default. The fear of stigma had the highest adjusted odds ratio, confirming that TB continues to be heavily stigmatized due to its association with HIV and the perception of TB as a “disease of poverty.” In Kalingalinga, a densely populated informal settlement, patients may fear judgment from neighbours or employers, leading them to hide their condition and skip clinic appointments. This has been echoed in Ghana and Nigeria where fear of disclosure remains a prominent cause of non-adherence [12,11]. Moreover, stigma not only discourages treatment seeking but may also isolate patients socially and emotionally, compounding depression and hopelessness factors known to negatively affect self-care behavior.

Substance abuse significantly increased the likelihood of default. Individuals dependent on alcohol or other substances often exhibit erratic health-seeking behaviors and may deprioritize structured routines like medication adherence. Substance abuse is also associated with increased financial hardship and mental health challenges, which can further complicate treatment. In low-resource settings like Zambia, limited access to rehabilitation or psychosocial support services leaves this group particularly vulnerable. Targeted interventions, including brief behavioral therapy and community-based addiction counselling, could improve outcomes in this high-risk subgroup.

Another critical patient-related factor was inadequate knowledge about TB. Misunderstandings about the curability of TB, treatment duration, and the potential for relapse can lead patients to abandon therapy prematurely, especially after symptomatic improvement. Similar gaps in knowledge were identified in Indonesia and Ethiopia, where the implementation of peer-led and pictorial education programs significantly improved adherence [13,9]. This highlights the need for simple, repeated health messages using multiple platforms such as radio, clinic posters, drama groups, and community health workers.

In terms of health system factors, the lack of counselling services was strongly associated with default. Effective counselling can enhance understanding, provide motivation, and serve as a platform to address stigma and misconceptions. However, in many Zambian primary health facilities, TB counselling is under-resourced or delegated to untrained staff due to human resource constraints. This finding aligns with research in Kenya and Malawi, which points to health worker shortages and poor communication skills as major deterrents to TB treatment adherence [14,15]. Investing in training for lay health workers and community-based adherence counsellors could bridge this critical gap.

Interruption of drug supply also emerged as a significant contributor to treatment default. Stock-outs not only disrupt medication intake but erode patient trust in the health system. Patients who experience stock-outs may become discouraged or perceive treatment as optional. Drug availability is a basic pillar of successful TB programs, and its unreliability reflects broader systemic issues in supply chain management. Strengthening drug procurement and inventory systems must be a policy priority, particularly in high-burden settings. Additionally, establishing buffer stocks at facility and district levels may prevent interruptions.

Distance to health facilities was another structural barrier. In Kalingalinga, patients living far from TB treatment centres were over three times more likely to default, often due to transport costs and time constraints. This burden is particularly heavy on daily wage earners who cannot afford to miss work or pay for repeated transportation. In similar peri-urban settings in Nigeria and Bangladesh, decentralized treatment models, home-based DOT, and digital adherence monitoring have shown promise in reducing geographic barriers [11,16]. Zambia could consider expanding community-based directly observed therapy through trained volunteers or deploying mobile TB clinics in hard-to-reach zones.

Overall, the findings indicate that improving TB treatment adherence requires multi-pronged strategies that go beyond individual behaviour change. Strengthening the health system to provide reliable, accessible, and patient-centred services is key. At the same time, targeted interventions addressing stigma, substance abuse, and health education must be integrated into TB control programs. Community engagement and the use of digital tools could enhance monitoring, education, and support, especially among high-risk groups such as men, the economically active population, and individuals with low literacy.

## Conclusion

The findings of this quantitative study highlight that tuberculosis treatment default in Kalingalinga Compound is influenced by a complex interplay of patient-level and health system factors. Key predictors of default included being male, aged between 30 and 49 years, having low educational attainment, fear of stigma, substance abuse, poor knowledge about TB, lack of counselling services, drug stock-outs, and long distances to treatment centres. These results suggest that default is not merely a result of patient unwillingness to adhere but often reflects structural and psychosocial challenges. To improve treatment adherence, TB control programs must prioritize interventions that address both individual behaviours and systemic barriers. Health education efforts should be intensified, stigma must be actively countered through community engagement, and substance abuse support should be integrated into TB care. Furthermore, ensuring consistent drug supply, expanding counselling services, and improving accessibility to treatment facilities are critical. Future research using complementary methodologies, including qualitative studies, could help uncover the nuanced experiences behind these patterns and inform more targeted interventions.

## Data Availability

All relevant data are within the manuscript and its Supporting Information files.

## Acknowledgments

The author extends sincere thanks to the University of Lusaka for academic support, and to the research supervisor for invaluable guidance throughout the study. Gratitude is also expressed to classmates and friends for their encouragement and support.

## Author Contributions

**Conceptualization:** Luyando Mainza, Chipampe Lombe

**Data curation:** Luyando Mainza

**Formal analysis:** Luyando Mainza, Chipampe Lombe

**Methodology:** Luyando Mainza, Chipampe Lombe

**Project administration:** Luyando Mainza, Chipampe Lombe

**Supervision:** Chipampe Lombe

**Validation:** Luyando Mainza, Chipampe Lombe

**Visualization:** Luyando Mainza, Chipampe Lombe

**Writing – original draft:** Luyando Mainza,

**Writing – review & editing:** Luyando Mainza, Chipampe Lombe

